# Optimal dosing of cefotaxime and desacetylcefotaxime for critically ill paediatric patients. Can we use microsampling?

**DOI:** 10.1101/2022.03.11.22272285

**Authors:** Yarmarly C. Guerra Valero, Tavey Dorofaeff, Mark G Coulthard, Louise Sparkes, Jeffrey Lipman, Steven C. Wallis, Jason A. Roberts, Suzanne L. Parker

## Abstract

**Objectives:** This study aimed to describe the population pharmacokinetics of cefotaxime and desacetylcefotaxime in critically ill paediatric patients and provide dosing recommendations. We also sought to evaluate the use of capillary microsampling to facilitate data-rich blood sampling.

**Methods:** Patients were recruited into a pharmacokinetic study, with cefotaxime and desacetylcefotaxime concentrations from plasma samples collected at 0, 0.5, 2, 4 and 6 h used to develop a population pharmacokinetic model using Pmetrics. Monte Carlo dosing simulations were tested using a range of estimated glomerular filtration rates (60, 100, 170 and 200 mL/min/1.73 m2) and body weights (4, 10, 15, 20 and 40 kg) to achieve PK/PD targets, including 100% ƒT>MIC with an MIC breakpoint of 1 mg/L.

**Results:** 36 patients (0.2 – 12 y) provided 160 conventional samples for inclusion in the model. The pharmacokinetics of cefotaxime and desacetylcefotaxime were best described using one-compartmental model with first-order elimination. The clearance and volume of distribution for cefotaxime were 12.8 L and 39.4 L/h, respectively. The clearance for desacetylcefotaxime was 10.5 L/h. Standard dosing of 50 mg/kg Q6h was only able to achieve the PK/PD target of 100% ƒT>MIC in patients > 10 kg and with impaired renal function or patients of 40 kg with normal renal function.

**Conclusions:** Dosing recommendations support the use of extended or continuous infusion to achieve cefotaxime exposure suitable for bacterial killing in critically ill paediatric patients, including those with severe or deep-seated infection. An external validation of capillary microsampling demonstrated skin-prick sampling can facilitate data-rich pharmacokinetic studies.

## Introduction

Severe infection can have long-term health consequences for paediatric patients, including impaired neurodevelopment and chronic disability ^1, 2^. Effective antimicrobial dosing is one of the cornerstones of care to ensure therapeutic success in the treatment of severe infection. However, critical illness can manifest as extreme physiological derangements and this has the potential to impact on drug exposure, leading to treatment failure and/or antimicrobial resistance ^3^.

Cefotaxime—a semisynthetic, third-generation cephalosporin—is one of the most prescribed antimicrobials used to treat severe infections in critically ill paediatric patients ^4-6^. Approximately 43% of cefotaxime is bound to plasma proteins and it exhibits good penetration into body fluids and tissues ^7, 8^. Cefotaxime is a hydrophilic drug with approximately 50 – 60% eliminated by the kidneys by glomerular filtration followed by tubular secretion ^9^. Cefotaxime is metabolised by enzymatic hydrolysis of the O-acetyl group by an acetyl esterase in the liver to a pharmacologically active metabolite, desacetylcefotaxime ^10, 11^. The metabolite is estimated as being between half to ten times less microbiologically active than the parent compound, cefotaxime ^8^. Optimal cefotaxime dosing regimens target concentrations above the minimum inhibitory concentration (MIC) throughout the dosing interval (pharmacokinetic/pharmacodynamic (PK/PD) ƒT>MIC), with targets of ≥60% ƒT>MIC and ≥100% ƒT>MIC for critically ill patients, and ≥100% ƒT>4xMIC for critically ill patients with severe or deep-seated infection. Cefotaxime can be used to treat infections caused by Gram-positive and Gram-negative organisms, including meningitis caused by *Escherichia coli, Neisseria meningitidis, Haemophilus influenzae*, and *Streptococcus pneumoniae* ^12^. Of the pathogens treated with cefotaxime, the reported MIC value according to the European Committee on Antimicrobial Susceptibility Testing (EUCAST) is 1 mg/L for meningitis and indications other than meningitis caused by *E. coli* ^13^. Additionally, an MIC non-species related breakpoint of 1 mg/L is commonly used for the treatment of a susceptible pathogen, with an MIC of ≥2 mg/L indicating a resistant pathogen ^13^. Current cefotaxime dosing regimens of 50 mg/kg every 6 h, with a maximum dose of 2 g (a total daily dose of up to 8 g), are commonly used for critically ill paediatric patients (>one month old of life) ^14-16^.

The primary aims of this study were (i) to describe the population pharmacokinetics of cefotaxime and desacetylcefotaxime in critically ill paediatric patients and to provide dosing recommendations for this special patient population and (ii) to describe the suitability of using capillary microsampling for blood sampling compared to samples collected from an indwelling arterial or venous cannula (conventional sampling) by performing an external validation.

## Patients and Methods

### Study Design

A prospective, open-label, pharmacokinetic study was conducted at the paediatric intensive care unit at the Queensland Children’s Hospital, Brisbane, Australia between March 2019 and September 2021. Critically ill patients between the ages of 1 month and 12 years and receiving intravenous cefotaxime, as prescribed by the treating physician, were included. Patients receiving extracorporeal membrane oxygenation, renal replacement therapy and peritoneal dialysis were excluded from this study. The research was conducted in accordance with the guidelines of the Declaration of Helsinki and approved by the Human Research & Ethics Committee of the Queensland Children’s Hospital (HREC/17/QRCH/45). Written informed consent was obtained from the parents or legal guardians prior to commencement of the study. Clinical characteristics were collected for patients including sex, age, height, weight, total bilirubin, haemoglobin, albumin, platelet count, white cell count, serum creatinine, alanine aminotransferase, aspartate aminotransferase, alkaline phosphatase, gamma-glutamyl transferase, prothrombin time, activated partial thromboplastin time, C-reactive protein, urinary creatinine, paediatric logistic organ dysfunction-2 (PELOD-2) score. For each patient, an estimated glomerular filtration rate (eGFR) was calculated using the bedside Schwartz equation (mL/min/1.73 m^2^) ^17, 18^.

### Conventional blood sampling and capillary microsampling

Paired blood samples using conventional sampling (from an arterial or venous line) and capillary microsamples ^19^ (from a finger or heel prick) were simultaneously collected at five pre-defined time points: prior to administration of the cefotaxime dose (time 0), and then after the end of infusion at approximately 0.5, 2, 4 and 6 h. For capillary microsamples, the patient’s finger was cleaned with alcohol and punctured using a lancet device (either Haemolance Plus ®, low flow 25G x 1.4mm or BD microtainer Quikheel Infant Lancet, 1mm x 2.5mm). The finger was gently massaged and held below the heart of the patient until approximately 50 µL of blood was collected into a heparinised plastic capillary tube. The capillary microsample was centrifuged at 2000 g for 10 minutes to obtain plasma. The capillary tube was then snipped with scissors to isolate the plasma. For conventional plasma samples, approximately 0.6 mL of blood was obtained and collected into a heparinised 1 mL vacuum tubes and centrifuged at 1500 g for 10 minutes to obtain plasma. After centrifugation, all plasma samples were transferred into screw-capped 2 mL polypropylene tubes and stored at -80 °C until analysis.

### Analysis of samples

Cefotaxime and desacetylcefotaxime concentrations were measured using a validated ultra high performance liquid chromatography tandem mass spectrometry (UHPLC-MS/MS) bioanalytical method ^20^ in accordance with the guidelines provided by the European Medicines Agency (EMA) ^21^ and the U.S. Food and Drug Administration (USFDA) ^22^. The linear concentration range was 0.5 – 500 mg/L and 0.2 – 10 mg/L for cefotaxime and desacetylcefotaxime, respectively. All intra-assay and inter-assay accuracy and precision were within 15% of acceptance criteria.

### Pharmacokinetic model

Pmetrics version 1.5.0 (Laboratory of Applied Pharmacokinetics and Bioinformatics, Los Angeles, CA, USA) in RStudio (version 0.99.9.3) as a wrapper for R (version 3.3.1), Xcode (version 2.6.2) and the Intel Parallel Studio Fortran Compiler XE 2017 was used to develop a population pharmacokinetic model. One-and two-compartment models were constructed using non-parametric adaptive grid (NPAG) algorithms with total plasma cefotaxime and desacetylcefotaxime concentrations. A stepwise approach was followed to build the model to establish (i) the structural base model, (ii) the best-fit error model, and (iii) development of a covariate model. Elimination from the central compartment and the rate of formation of the metabolite were modelled as first-order processes, and rate of formation of the metabolite was also tested for Michaelis-Menten kinetics. Lambda (additive) and gamma (multiplicative) error models were evaluated using a polynomial equation for standard deviation as a function of observed concentration with observation weighting performed as error = SD.gamma or error = (SD^2^ + lambda^2^)^0.5^.

### Pharmacokinetic model evaluation

The coefficient of determination (*R*^*2*^) was used to select the final model. The visual predictive check plot, log-likelihood ratio (−2*LL) and Akaike information criterion were used to compare different models. The bias was measured using the mean weighted predicted – observed error. Imprecision was measured by using bias-adjusted and the mean weighted squared predicted-observed error. The percentage of shrinkage was measured using the total variation in the probability of each model.

### External validation

An external validation was performed to describe the correlation between the measurement of cefotaxime concentrations obtained from capillary microsamples compared to the concentrations obtained using conventional sampling. For the external validation, the model developed using conventional sampling was used as a prior and Bayesian posterior simulations was calculated for each subject. A linear regression, the goodness of fit and the coefficient of determination were used to assess the correlation between the observed and predicted concentrations. Prediction errors were evaluated to describe bias (calculated as mean weighted prediction error, MWPE) and precision (Root Mean Square Predication error, RMSE) using Pmetrics. Bland-Altman plots were used to visually inspect the observed (model simulated conventional sampling) and predicted (capillary microsampling) cefotaxime and desacetylcefotaxime concentrations for systematic bias.

### Dosing simulations

Cefotaxime dosing regimens administered as a bolus dose every 4 or 6 h, as a 2 or 3 h extended infusion (EI), or as a continuous infusion (CI) across a range of eGFR (60, 100, 170 and 200 mL/min/1.73 m^2^) and a range of body weights (4, 10, 15, 20 and 40 kg) were evaluated using Monte Carlo simulations (n = 1000) in Pmetrics. Cefotaxime protein binding at 40% was used to calculate the probability of target attainment (PTA) ^23^. For each dosing regimen, the PTA was calculated as the percentage of patients achieving a ≥60% ƒT>MIC, ≥100% ƒT>MIC or ≥100% ƒT>4xMIC with MIC non-species related breakpoint of 1 mg/L ^13^, targeting success at 90%.

## Results

A total of 36 critically ill paediatric patients (median age: 30.4 months [interquartile range age: 8.2 – 65.8 months]) with 160 conventional samples were included in the model development. Eleven plasma samples were excluded from the analysis as the concentrations were below the lower limit of quantification for both cefotaxime and desacetylcefotaxime. Cefotaxime was administered as an intermittent infusion in 34 patients over the mean ± SD duration of 0.20 ± 0.22 h; two patients received an extended infusion over the mean ± SD duration of 4.5 ± 1.1 h.

From the total study cohort, 58% of the patients (n = 21) had eGFR > 130 mL/min/1.73 m^2^, 33% (n = 12) of the patients had renal function with an eGFR ranging between 80 and 130 mL/min/1.73 m^2^, while 8% (n = 3) of the patients had eGFR values < 80 mL/min/1.73 m^2^. Of the patients recruited, 39% (n = 14) weighed less than 10 kg, 47% of the patients weighed between 10 and 30 kg (n = 17) and 14% of the patients (n = 5) had a body weight above 30 kg. The baseline characteristics and clinical information from the patients are presented in Table 1.

**Table 1:** Clinical characteristics and information

| Demographic data | Median* (n = 36) |
| --- | --- |
| Sex, n (%) | Female 14 (39%)<br>Male 22 (61%) |
| Age (months) | 30.4 (8.2 – 65.8) |
| Height (cm) | 86.5 (68.5 – 109) |
| Weight (kg) | 11.7 (8.1 – 18.2) |
| Body surface area (m<sup>2</sup>) | 0.5 (0.4 – 0.7) |
| Albumin (g/L) | 30 (24 – 33) |
| Bilirubin (μmol/L) | 6.0 (3.5 – 9.5) |
| Haemoglobin (g/L) | 106 (97 – 116) |
| Platelet count (x 10<sup>9</sup>/L) | 251 (189 – 306) |
| Serum Creatinine (mL/min) | 24.0 (17.5 – 28.5) |
| eGFR (Indexed to BSA 1.73, calculated using bedside Schwartz equation; mL/min/1.73 m<sup>2</sup>) | 143 (109 – 259) |
| Illness severity score PELOD-2 score (on admission to the ICU) | 4 (2 – 6) |
| Cefotaxime IV dose (mg/kg)** | 605 (404 – 910) |

\*Data displayed as mean with IQR (Q1 – Q3) or n (%) as appropriate. PELOD-2 score, Paediatric Logistic Organ Dysfunction; eGFR: estimated glomerular filtration rate.
\*\* Data displayed as mean (minimum – maximum)

Plasma-concentration data were best described using a one-compartmental model with first-order elimination for both cefotaxime and desacetylcefotaxime. For the model, empiric inclusion of weight normalised to 70 kg with allometric scaling (0.75) on clearance and linear scaling on volume of distribution was used on cefotaxime. As the volume of distribution for desacetylcefotaxime could not be estimated, it was assumed to be equal to the volume of distribution of cefotaxime ^24^. The inclusion of the patient population mean-adjusted eGFR (eGFR/150) was accepted as a covariate on cefotaxime clearance (CL1) as it resulted in a decrease in log-likelihood of 26.0. The inclusion of normalised body surface area (BSA/1.73 m^2^) as a covariate on desacetylcefotaxime clearance decreased the log-likelihood by 7.0. The goodness-of-fit of the final models were confirmed with the diagnostic plots shown in Figure 1. The final Pmetrics model is provided in the supplementary material (Table S1).

**Figure 1:**
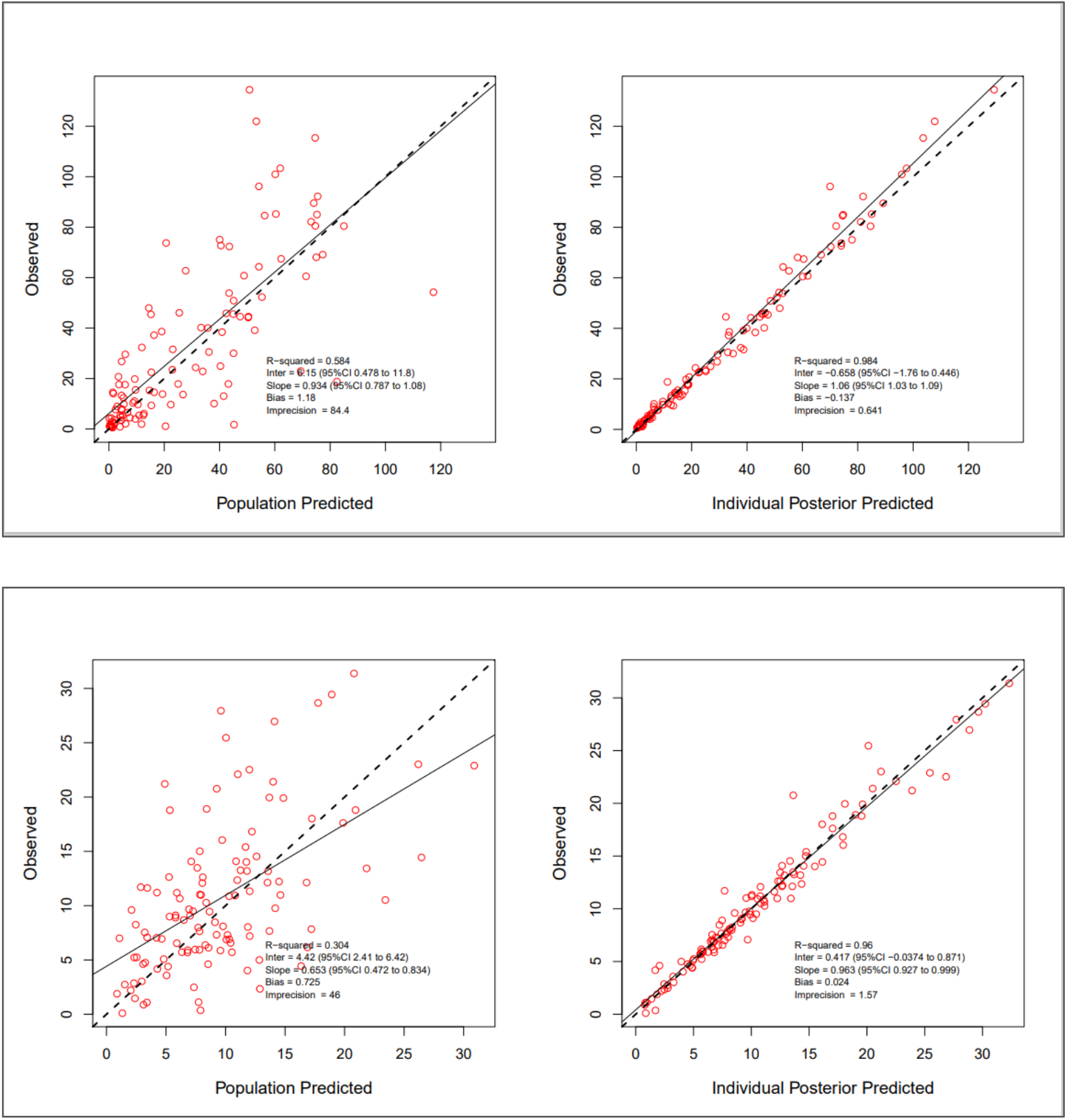
Diagnostic plots for the final covariate model for plasma concentrations (mg/L) of cefotaxime (top) and desacetylcefotaxime (bottom).

The primary pharmacokinetic parameters are summarised in Table 2 and the visual predicted check plots for cefotaxime and desacetylcefotaxime are provided in Figure 2. Based on the visual predictive check, 94.7% of the observations for cefotaxime and 96.6% of the observations for desacetylcefotaxime were within the 5^th^ and 95^th^ of simulated percentiles. Individual plots are presented in the supplementary material (Figures S1 – S4). Probability of target attainment for cefotaxime based on the PK/PD targets of ≥60% ƒT>MIC, ≥100% ƒT>MIC, and ≥100% ƒT>4xMIC are presented across the range of patient weights and eGFR in Tables 3A–C.

**Table 2:**
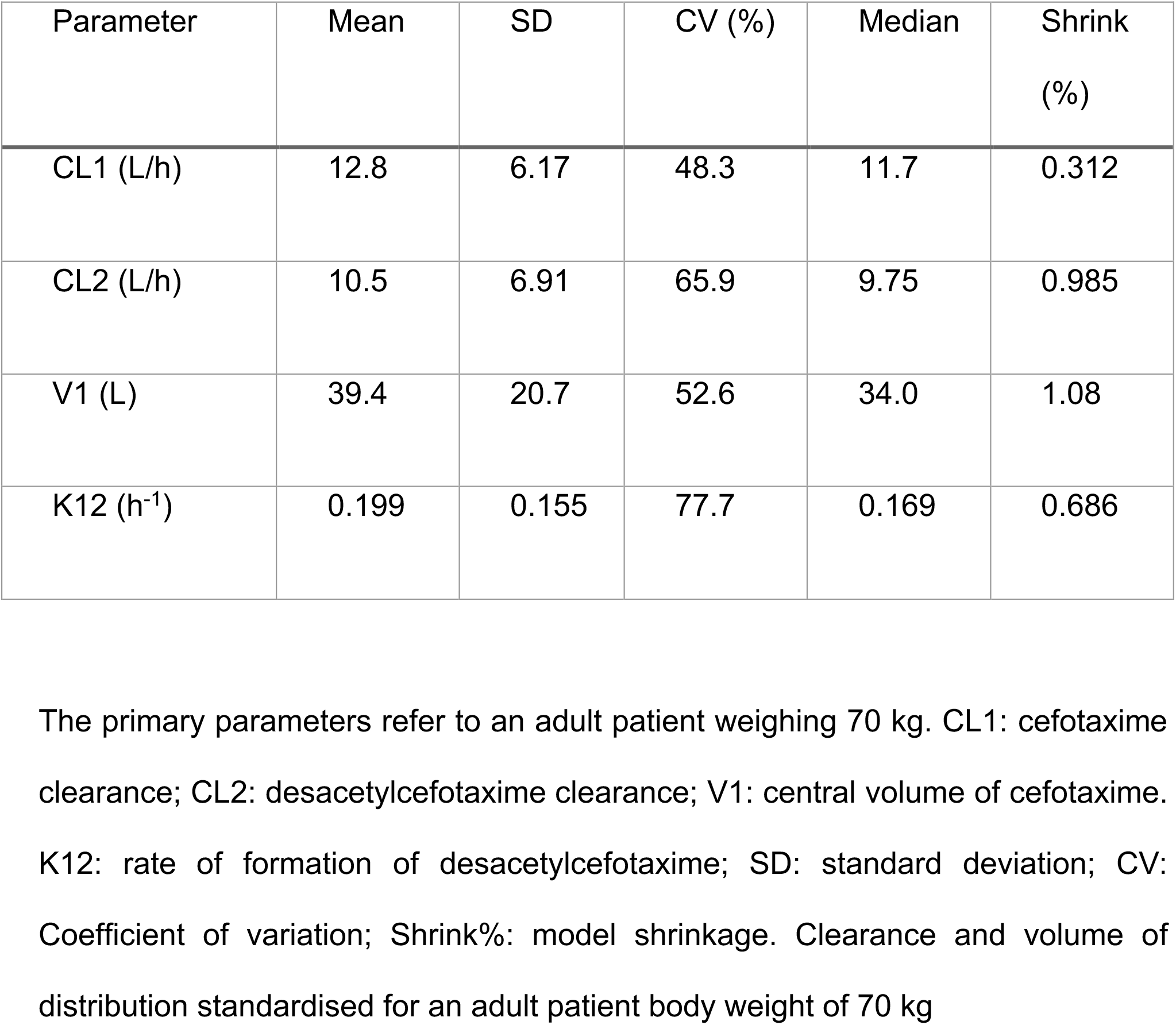
Population pharmacokinetic primary parameters of cefotaxime and desacetylcefotaxime concentrations of critically ill paediatric patients

| Parameter | Mean | SD | CV (%) | Median | Shrink (%) |
| --- | --- | --- | --- | --- | --- |
| CL1 (L/h) | 12.8 | 6.17 | 48.3 | 11.7 | 0.312 |
| CL2 (L/h) | 10.5 | 6.91 | 65.9 | 9.75 | 0.985 |
| V1 (L) | 39.4 | 20.7 | 52.6 | 34.0 | 1.08 |
| K12 (h<sup>-1</sup>) | 0.199 | 0.155 | 77.7 | 0.169 | 0.686 |

The primary parameters refer to an adult patient weighing 70 kg. CL1: cefotaxime clearance; CL2: desacetylcefotaxime clearance; V1: central volume of cefotaxime. K12: rate of formation of desacetylcefotaxime; SD: standard deviation; CV: Coefficient of variation; Shrink%: model shrinkage. Clearance and volume of distribution standardised for an adult patient body weight of 70 kg

**Table 3A:**
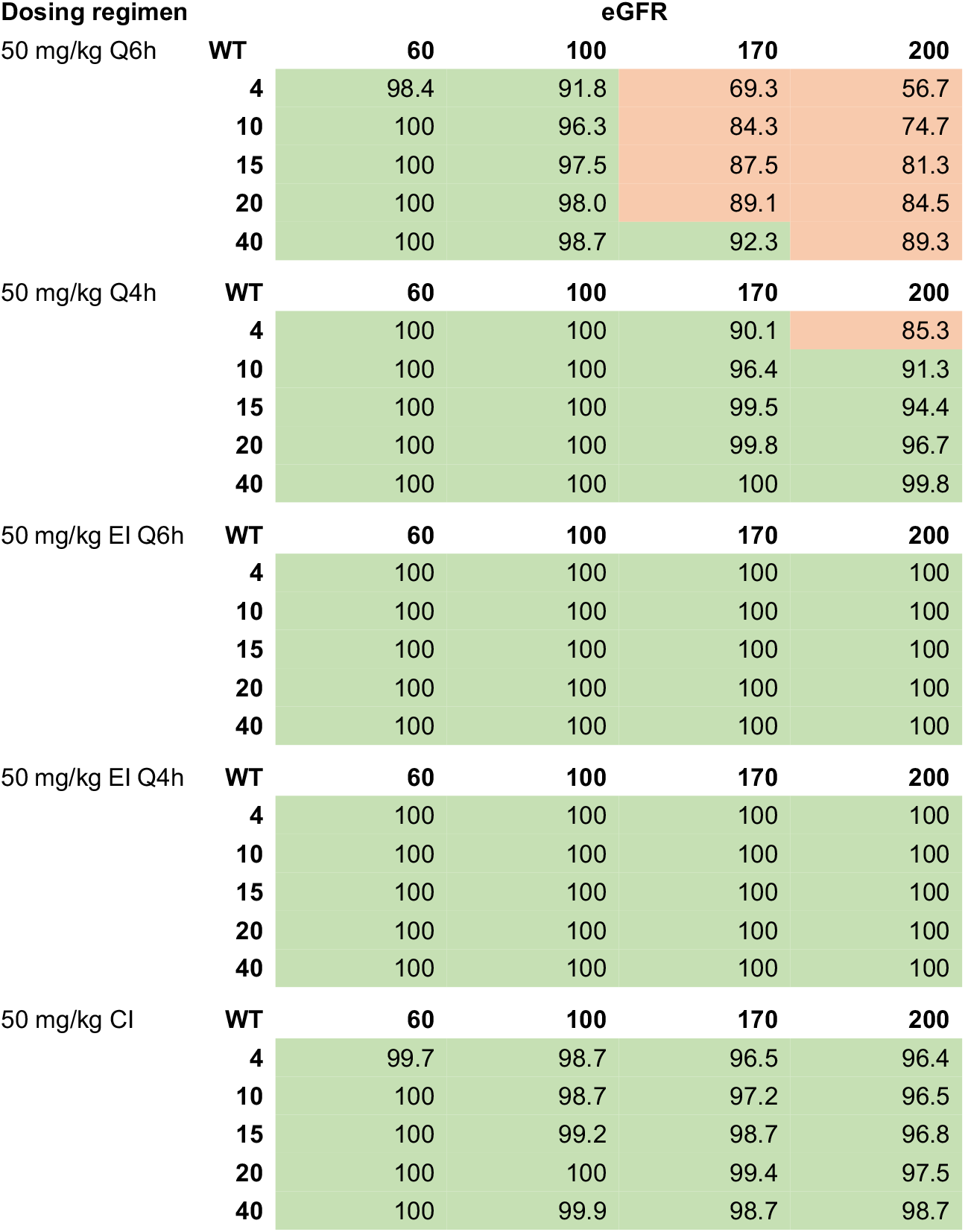
Dose simulations (with probability of target attainment results, %) for pathogens susceptible to cefotaxime (MIC target of 1 mg/L) to achieve a PK/PD target of 60% *f*T/MIC

**Table 3B:** Dose simulations (with probability of target attainment results, %) for pathogens susceptible to cefotaxime (MIC target of 1 mg/L) to achieve a PK/PD target of 100% *f*T/MIC

| Dosing regimen |  | eGFR |  |  |  |
| --- | --- | --- | --- | --- | --- |
|  | WT | 60 | 100 | 170 | 200 |
| 50 mg/kg Q6h | 4 | 89.7 | 55.8 | 29.1 | 18.4 |
|  | 10 | 93.0 | 74.0 | 41.6 | 33.0 |
|  | 15 | 94.9 | 81.3 | 46.5 | 38.2 |
|  | 20 | 95.9 | 86.4 | 49.2 | 42.2 |
|  | 40 | 97.2 | 91.4 | 58.6 | 49.7 |
| 50 mg/kg Q4h | WT | 60 | 100 | 170 | 200 |
|  | 4 | 97.2 | 88.1 | 59.5 | 51.6 |
|  | 10 | 98.1 | 91.4 | 76.0 | 64.2 |
|  | 15 | 99.3 | 94.9 | 81.7 | 71.2 |
|  | 20 | 99.8 | 96.5 | 84.7 | 76.3 |
|  | 40 | 100 | 97.5 | 88.8 | 85.0 |
| 50 mg/kg EI Q6h | WT | 60 | 100 | 170 | 200 |
|  | 4 | 97.3 | 89.0 | 56.3 | 47.6 |
|  | 10 | 97.6 | 92.8 | 75.9 | 60.5 |
|  | 15 | 97.9 | 95.4 | 81.9 | 69.1 |
|  | 20 | 98.2 | 96.2 | 84.7 | 76.2 |
|  | 40 | 99.1 | 97.4 | 90.0 | 85.4 |
| 50 mg/kg EI Q4h | WT | 60 | 100 | 170 | 200 |
|  | 4 | 100 | 99.7 | 85.8 | 79.9 |
|  | 10 | 100 | 100 | 91.5 | 87.9 |
|  | 15 | 100 | 100 | 94.8 | 90.4 |
|  | 20 | 100 | 100 | 96.7 | 91.5 |
|  | 40 | 100 | 100 | 99.8 | 96.9 |
| 50 mg/kg CI | WT | 60 | 100 | 170 | 200 |
|  | 4 | 99.7 | 98.7 | 96.5 | 96.4 |
|  | 10 | 100 | 98.7 | 97.2 | 96.5 |
|  | 15 | 100 | 99.2 | 98.7 | 96.8 |
|  | 20 | 100 | 100 | 99.4 | 97.5 |
|  | 40 | 100 | 99.9 | 98.7 | 98.7 |

**Table 3C:**
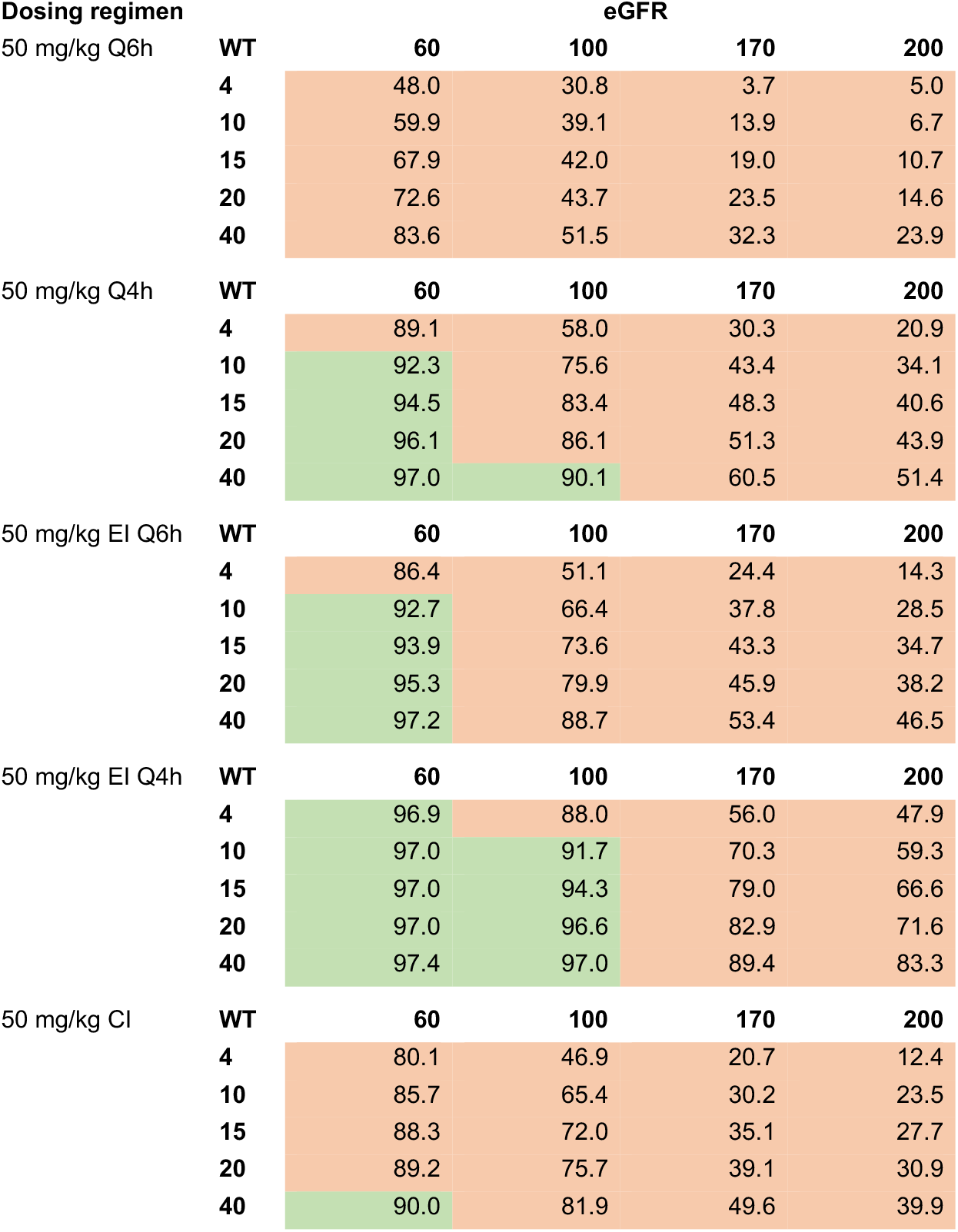
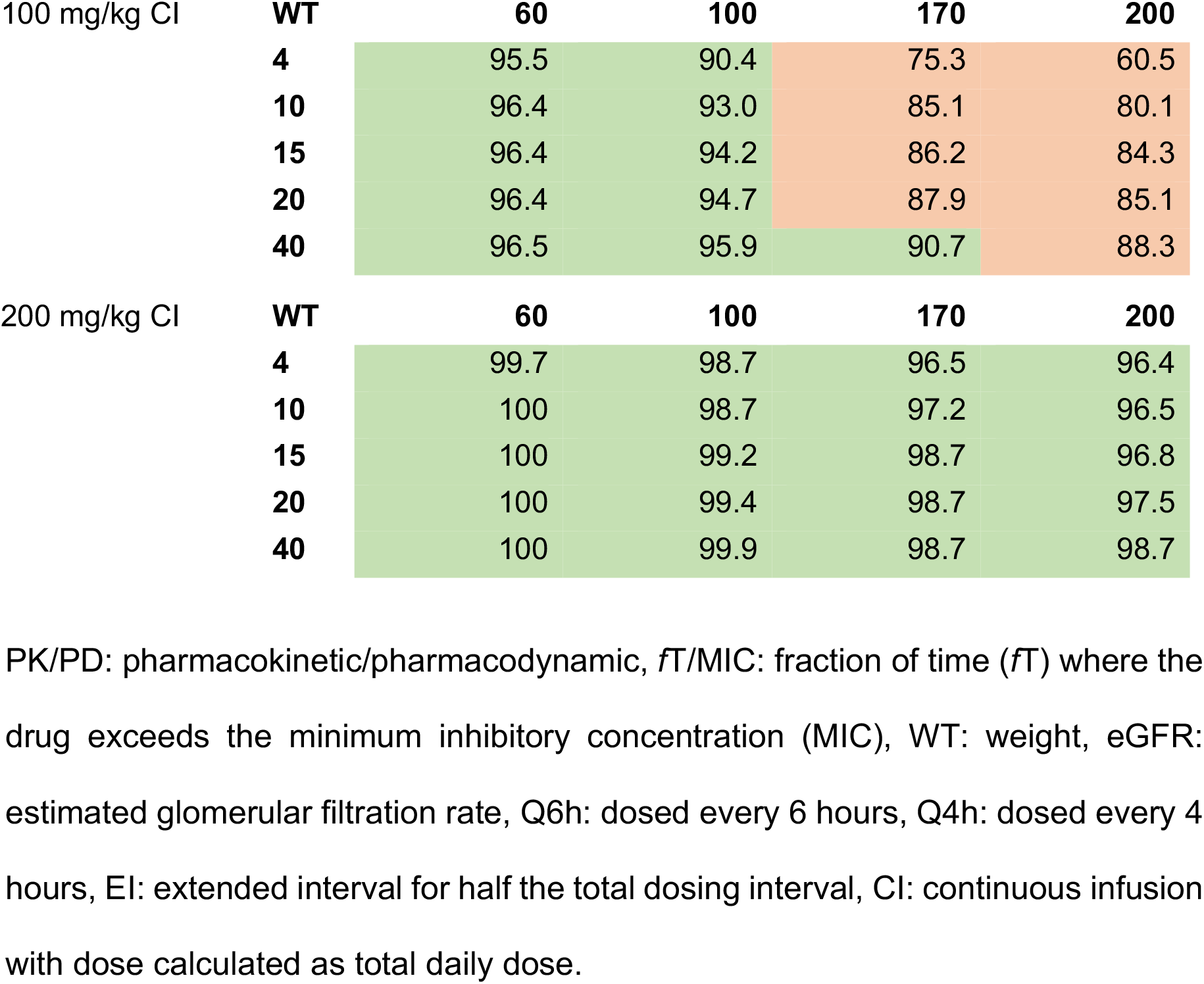
Dose simulations (with probability of target attainment results, %) for pathogens susceptible to cefotaxime (MIC target of 1 mg/L) to achieve a PK/PD target of 100% *f*T/4xMIC

| Dosing regimen |  | eGFR |  |  |  |
| --- | --- | --- | --- | --- | --- |
|  | WT | 60 | 100 | 170 | 200 |
| 50 mg/kg Q6h | WT |  |  |  |  |
|  | 4 | 48.0 | 30.8 | 3.7 | 5.0 |
|  | 10 | 59.9 | 39.1 | 13.9 | 6.7 |
|  | 15 | 67.9 | 42.0 | 19.0 | 10.7 |
|  | 20 | 72.6 | 43.7 | 23.5 | 14.6 |
|  | 40 | 83.6 | 51.5 | 32.3 | 23.9 |
| 50 mg/kg Q4h | WT |  |  |  |  |
|  | 4 | 89.1 | 58.0 | 30.3 | 20.9 |
|  | 10 | 92.3 | 75.6 | 43.4 | 34.1 |
|  | 15 | 94.5 | 83.4 | 48.3 | 40.6 |
|  | 20 | 96.1 | 86.1 | 51.3 | 43.9 |
|  | 40 | 97.0 | 90.1 | 60.5 | 51.4 |
| 50 mg/kg EI Q6h | WT |  |  |  |  |
|  | 4 | 86.4 | 51.1 | 24.4 | 14.3 |
|  | 10 | 92.7 | 66.4 | 37.8 | 28.5 |
|  | 15 | 93.9 | 73.6 | 43.3 | 34.7 |
|  | 20 | 95.3 | 79.9 | 45.9 | 38.2 |
|  | 40 | 97.2 | 88.7 | 53.4 | 46.5 |
| 50 mg/kg EI Q4h | WT |  |  |  |  |
|  | 4 | 96.9 | 88.0 | 56.0 | 47.9 |
|  | 10 | 97.0 | 91.7 | 70.3 | 59.3 |
|  | 15 | 97.0 | 94.3 | 79.0 | 66.6 |
|  | 20 | 97.0 | 96.6 | 82.9 | 71.6 |
|  | 40 | 97.4 | 97.0 | 89.4 | 83.3 |
| 50 mg/kg CI | WT |  |  |  |  |
|  | 4 | 80.1 | 46.9 | 20.7 | 12.4 |
|  | 10 | 85.7 | 65.4 | 30.2 | 23.5 |
|  | 15 | 88.3 | 72.0 | 35.1 | 27.7 |
|  | 20 | 89.2 | 75.7 | 39.1 | 30.9 |
|  | 40 | 90.0 | 81.9 | 49.6 | 39.9 |
| 100 mg/kg CI | WT | 60 | 100 | 170 | 200 |
|  | 4 | 95.5 | 90.4 | 75.3 | 60.5 |
|  | 10 | 96.4 | 93.0 | 85.1 | 80.1 |
|  | 15 | 96.4 | 94.2 | 86.2 | 84.3 |
|  | 20 | 96.4 | 94.7 | 87.9 | 85.1 |
|  | 40 | 96.5 | 95.9 | 90.7 | 88.3 |
| 200 mg/kg CI | WT | 60 | 100 | 170 | 200 |
|  | 4 | 99.7 | 98.7 | 96.5 | 96.4 |
|  | 10 | 100 | 98.7 | 97.2 | 96.5 |
|  | 15 | 100 | 99.2 | 98.7 | 96.8 |
|  | 20 | 100 | 99.4 | 98.7 | 97.5 |
|  | 40 | 100 | 99.9 | 98.7 | 98.7 |

PK/PD: pharmacokinetic/pharmacodynamic, $fT/MIC$ : fraction of time ( $fT$ ) where the drug exceeds the minimum inhibitory concentration (MIC), WT: weight, eGFR: estimated glomerular filtration rate, Q6h: dosed every 6 hours, Q4h: dosed every 4 hours, EI: extended interval for half the total dosing interval, CI: continuous infusion with dose calculated as total daily dose.

**Figure 2:**
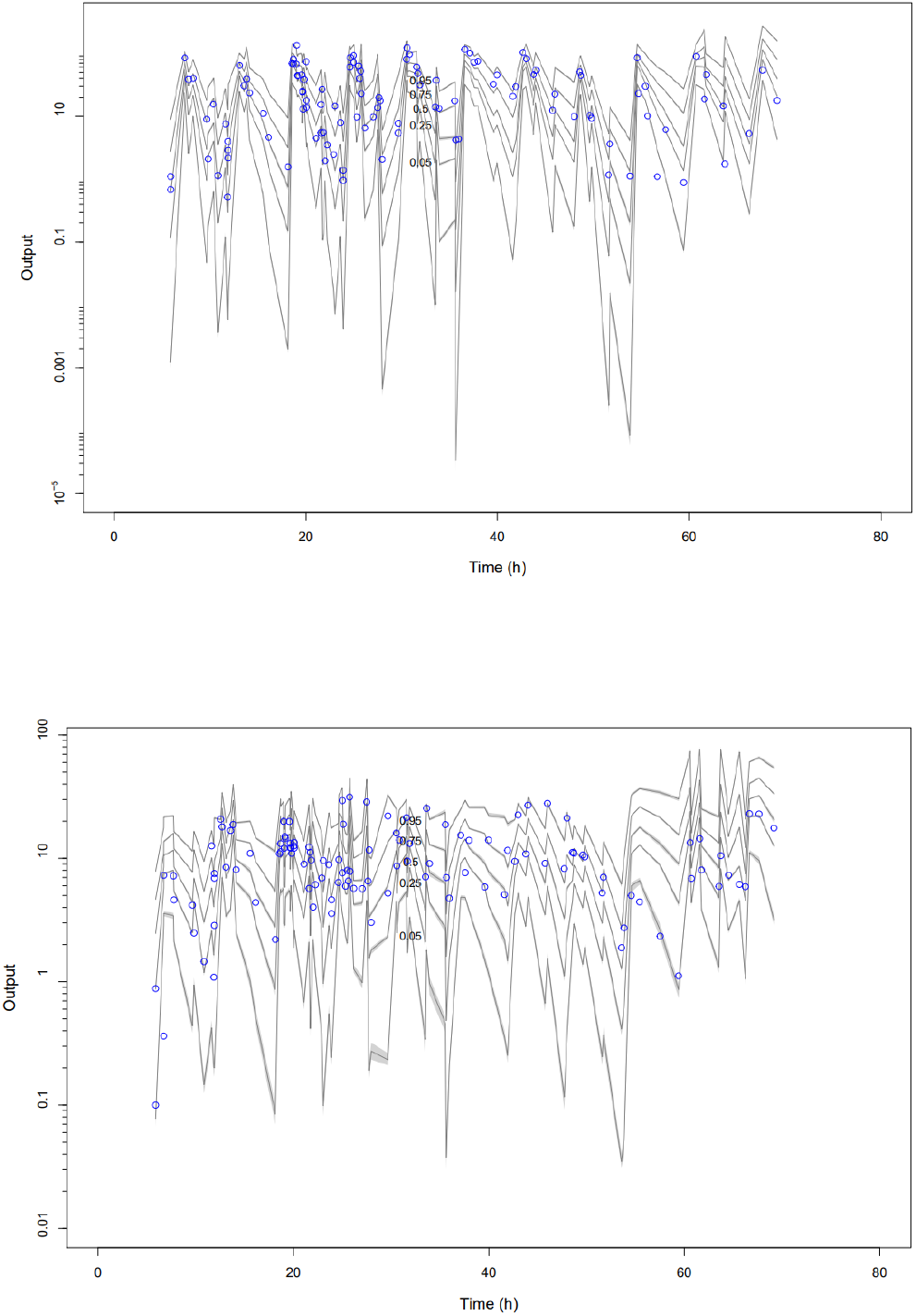
Visual predictive check for the final population pharmacokinetic model of cefotaxime (top) and desacetylcefotaxime (bottom). Observed data are represented by open circles. Lines represent the confidence intervals of the 5th, 25th, 50th, 75th and 95th percentiles of the simulated plasma concentrations. The y-axis is presented using a logarithmic scale.

The results of the external validation found for cefotaxime there was a bias (MWPE) of -0.137 mg/L (P = 0.1129, different than 0) and a precision (RMSE) of 14.6% and for desacetylcefotaxime there was a bias (MWPE) of -0.024 mg/L (P = 0.0967, different than 0) and a precision (RMSE) of 12.9%, when comparing the observed concentrations (capillary microsampling) to the model-predicted concentrations (using conventional sampling). A linear regression of the predicted versus observed cefotaxime and desacetylcefotaxime concentrations are presented in Figure 3. The 95% confidence intervals (95%CI) for the intercept for cefotaxime and desacetylcefotaxime are -1.8 to 0.45 and -0.04 to 0.87 mg/L, respectively and the slope of the regression line is close to 1 for both cefotaxime (95%CI 1.03 to 1.09) and desacetylcefotaxime (95%CI 0.927 to 0.999). The regression line crosses the line of equality for both cefotaxime and desacetylcefotaxime. The Bland-Altman weighted residual plots presented in Figure 4.

**Figure 3:**
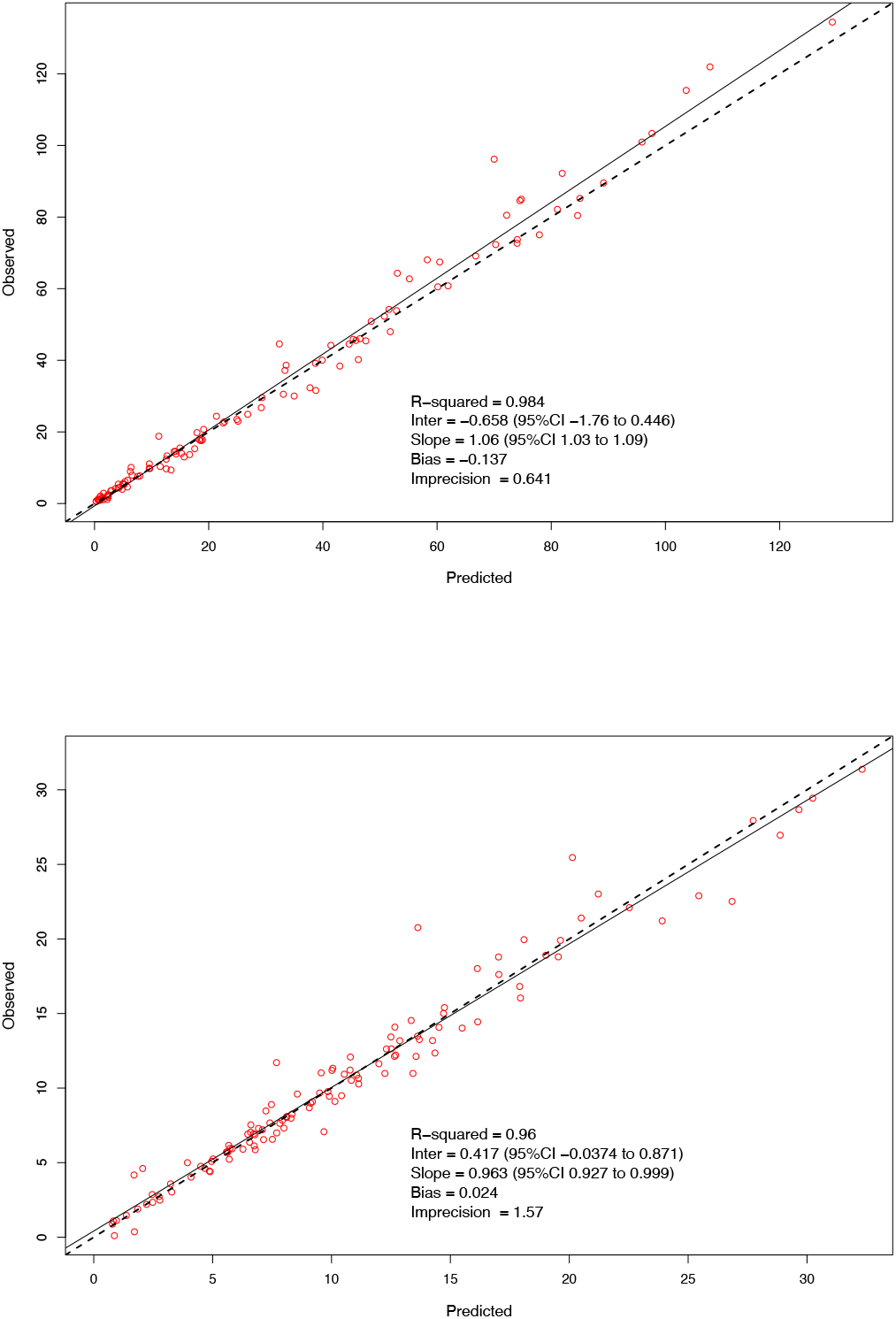
External validation linear regression plots of cefotaxime (top) and desacetylcefotaxime (bottom) comparing observed concentrations (capillary microsampling) with model-predicted concentrations (using conventional sampling).

**Figure 4:**
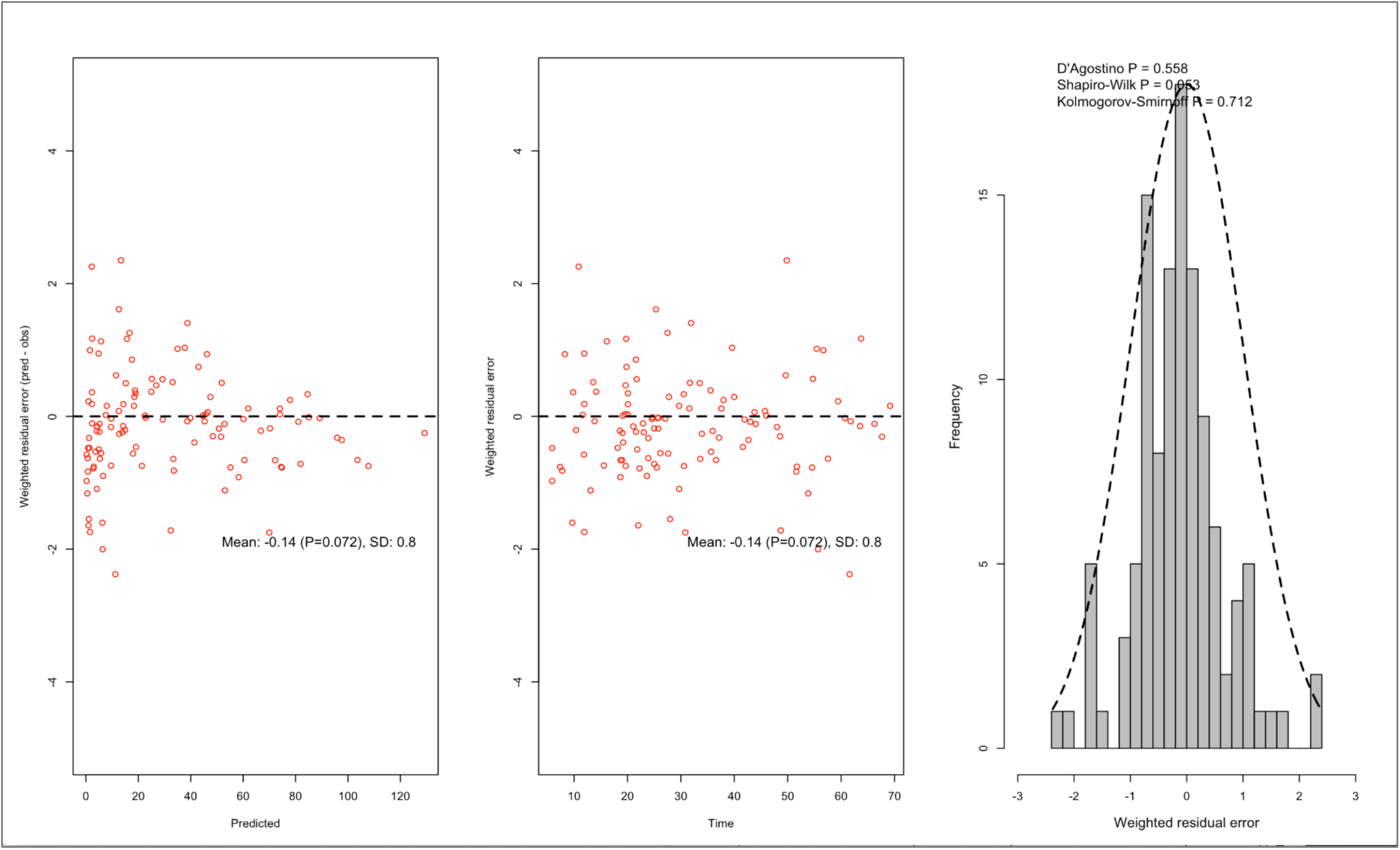
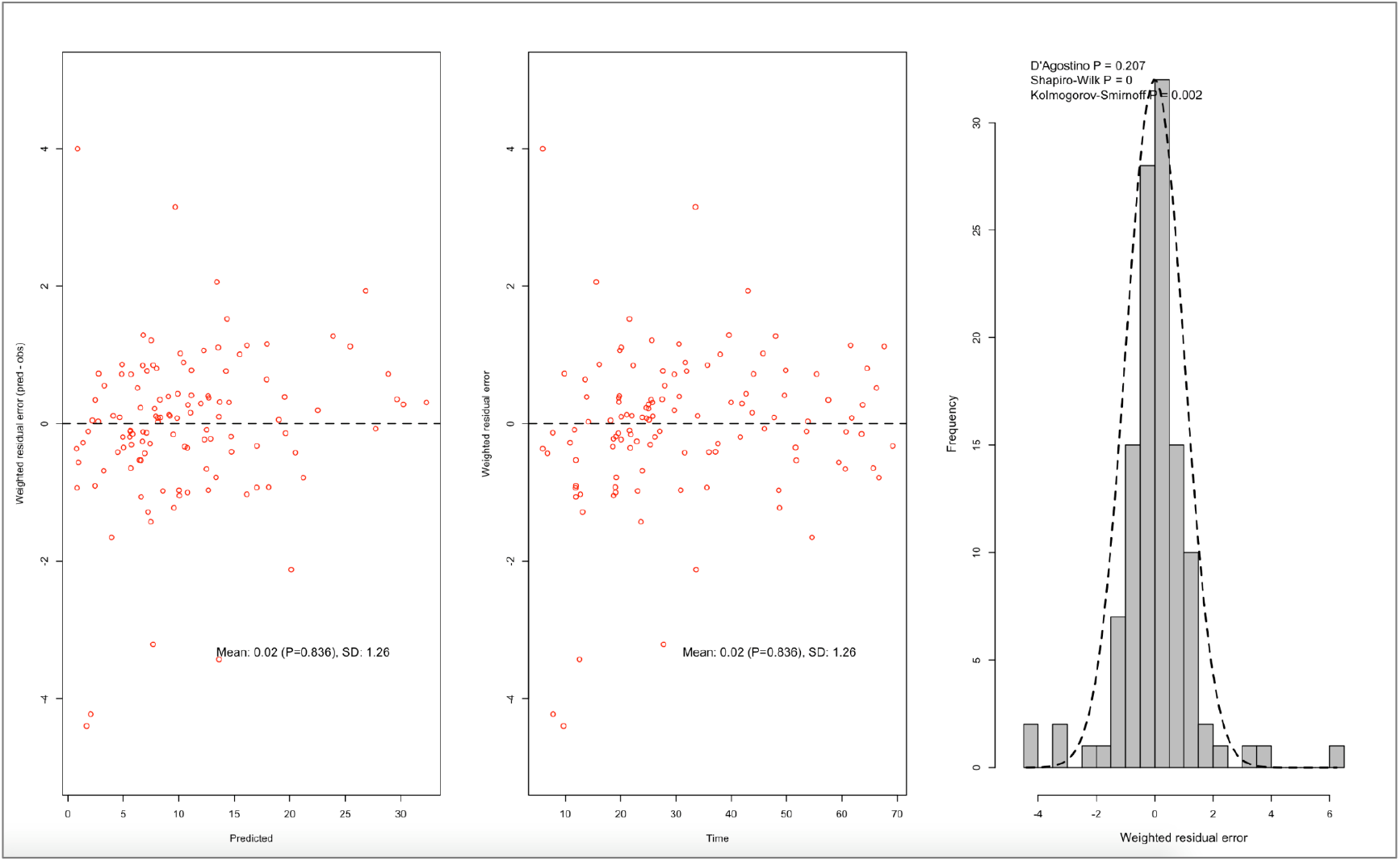
Bland-Altman weighted residual plots for external validation of cefotaxime (top) and desacetylcefotaxime (bottom) comparing observed concentrations (capillary microsampling) with model-predicted concentrations (using conventional sampling).

## Discussion

This study enhances our understanding of cefotaxime and, its active metabolite, desacetylcefotaxime pharmacokinetics and optimised dosing in critically ill paediatric patients ^24-26^. Through the use of rich blood sampling in paediatric patients to build the pharmacokinetic profiles (n = 5 samples/patient, range 3 to 5). Based on this study design, we have developed a model that supports the inclusion of eGFR on the clearance of cefotaxime and body surface area on the clearance of desacetylcefotaxime. Additionally, our blood sampling strategy has demonstrated that the application of capillary microsampling to obtain blood from a finger or heel prick correlates with concentrations obtained using conventional sampling techniques.

A one-compartmental model with first order elimination best fitted our data to describe the pharmacokinetics of cefotaxime and desacetylcefotaxime. In paediatric patients only one other study has used a population pharmacokinetic approach, which used a similar approach applied here, including setting the volume of distribution of desacetylcefotaxime to equal that of cefotaxime ^24^. Cefotaxime clearance was similar to that previously reported in critically ill paediatric patients, where clearance ranged from 6.9 to 13.7 L/h ^24-26^. All studies report variable cefotaxime clearance in critically ill patients. Desacetylcefotaxime clearance was higher in our study, compared to the study by Beranger et al, which reported a clearance of 4.2 L/h. Both studies had patients with a similar median renal function, so this may be a result of the increased definition allowed through the use of 2-4 samples per patient during the elimination phase in our study. Both studies have found the clearance of the active metabolite was highly variable. Cefotaxime and desacetylcefotaxime are eliminated by the kidneys and estimated creatinine clearance (using eGFR) was able to be included on the clearance of cefotaxime. This finding concords with data from critically ill adult patients that have shown clearance to be proportional to estimated creatinine clearance ^27^. No other studies have found an association between body surface area and clearance of desacetylcefotaxime, although a relationship between body surface area and liver volume has been recently identified in children ^28^ and this may account for the relationship identified for the metabolite in our study.

The volume of distribution for cefotaxime was variable in our patient cohort, but similar to other studies in critically ill paediatric patients, which have reported it ranging from 21.4 to 96 L ^24, 26^. Studies in critically ill patients have found the volume of distribution of hydrophilic antimicrobials, such as cefotaxime, can be highly altered due to a ‘third spacing’ phenomenon uneven distribution of body fluids ^29^. This may occur in patients suffering from capillary leak syndrome caused by severe sepsis or critically ill patients requiring extensive fluid resuscitation ^30^.

Based on a PK/PD target of ≥100% ƒT>MIC, with a non-species related breakpoint MIC of 1 mg/L for susceptible organisms ^31^, 14% of patients (n = 5) failed to achieve a target of 1 mg/L for cefotaxime and 39% of patients (n = 14) failed to achieve a target of 4 mg/L across the dosing interval. In our study cohort, 58% of patients had augmented renal clearance (defined as eGFR values above 130 mL/min/1.73 m^2^) ^32^. This result concords with other studies of both critically ill adults and paediatric patients 32-34.

Dosing simulations, using a range of weights and eGFR, support the use of shorter dosing intervals to achieve a PK/PD target of ≥60% ƒT>MIC. For critically ill paediatric patients with normal or impaired renal function, to achieve a PK/PD target of ≥100% ƒT>MIC a 4-hourly dosing interval or an extended infusion with a 6-hourly interval was able to provide sufficient cefotaxime coverage (using) for most patient weight and eGFR ranges. However, critically ill patients with augmented renal clearance, or neonatal patients with normal renal clearance required both a 4-hourly dosing interval combined with a 2-hour extended infusion to achieve the PK/PD target of ≥100% ƒT>MIC. More aggressive PK/PD targets (≥100% ƒT>4xMIC) that may be suitable for critically ill patients with severe or deep-seated infection, were not achieved using standard dosing of 50 mg/kg every 6 h. For critically ill paediatric patients with normal or impaired renal function a 4-hourly dosing interval combined with a 2-hour extended infusion achieved target in all patient weight ranges, except for neonatal patients with normal renal function. For critically ill patients with augmented renal clearance or neonatal patients with normal renal clearance, a continuous infusion with a total daily dose of 100 – 200 mg/kg was required to achieve this PK/PD target. Previous studies have demonstrated the challenge of achieving effective PK/PD targets for cefotaxime ^24^ and other beta-lactam antimicrobials ^35-38^ in critically ill paediatric patients with higher eGFR. The study by Beranger *et al*, targeting ≥100% ƒT>MIC and ≥100% ƒT>4xMIC for pathogens with an MIC of 0.5 mg/L, recommended the use of continuous infusion to achieve PK/PD targets in a similar patient population ^24^.

From the external validation, there is no systematic bias evident when comparing the concentration results of cefotaxime or desacetylcefotaxime obtained by conventional sampling to samples obtained by finger or heel prick using capillary microsampling. While the Bland Altman plots of weighted residual error over the predicted concentration range show a greater imprecision at low cefotaxime and desacetylcefotaxime concentrations, the histograms show that overall, there is a normal distribution of bias for both cefotaxime and desacetylcefotaxime across the predicted concentration range.

This study has several limitations. We measured total cefotaxime and desacetylcefotaxime concentrations in plasma samples and have not quantified the unbound concentrations. A consequence of this is that we have been unable to calculate protein binding for our patients and have set cefotaxime protein binding to 40% for the purpose of performing the probability of target attainment calculations and this may impact on the accuracy of the resultant dosing recommendations. Additionally, we did not collect and isolate the pathogens that caused the infections in the patients enrolled in the study and have therefore applied the pharmacokinetic-pharmacodynamic non-species related breakpoints from the European Committee on Antimicrobial Susceptibility Testing (EUCAST^31^) as targets to derive suitable dosing recommendations.

The strengths of this study are that it is the first pharmacokinetic study describing cefotaxime and desacetylcefotaxime in critically ill paediatric patients using rich-sampling for blood collection. Additionally, we demonstrate that the use of capillary microsampling can be used perform pharmacokinetic studies and there is the potential for this to facilitate more studies in neonatal and paediatric patients ^39^.

Standard dosing of 50 mg/kg every 6h was only able to achieve the PK/PD target commonly used in intensive care of 100% ƒT>MIC in patients > 10kg and with impaired renal function or patients of 40kg with normal renal function. Dosing recommendations support the use of shorter intervals or extended or continuous infusion to achieve cefotaxime exposure suitable for bacterial killing in critically ill paediatric patients, including patients with severe or deep-seated infection. Capillary microsampling for blood collection was externally validated and demonstrated the application of a finger/heel prick sample can facilitate data-rich pharmacokinetic studies.

## Data Availability

All data produced in the present study are available upon reasonable request to the authors

## Acknowledgements

The authors would like to acknowledge the critically ill children who participated in the study, their parents or legal guardians, the research nurses of the PICU at the Queensland Children’s Hospital, Brisbane Australia for their support and assistance with sample collection and other relevant tasks for this study. The authors also acknowledge the assistance of the staff of the Mass Spectrometry Facility of UQ Centre for Clinical Research, Brisbane, Australia.

## Funding

The study was funded by a grant from the Children’s Hospital Foundation, Queensland. Y Guerra Valero is a recipient of a Research Training Scholarship from The University of Queensland. SL Parker is a recipient of an Early Career Research Fellowship from the Australian National Health and Medical Research Council (APP1142757). JA Roberts is a recipient of an Australian National Health and Medical Research Council Fellowship (APP1048652).

## Transparency declarations

None to declare

**Table S1:**
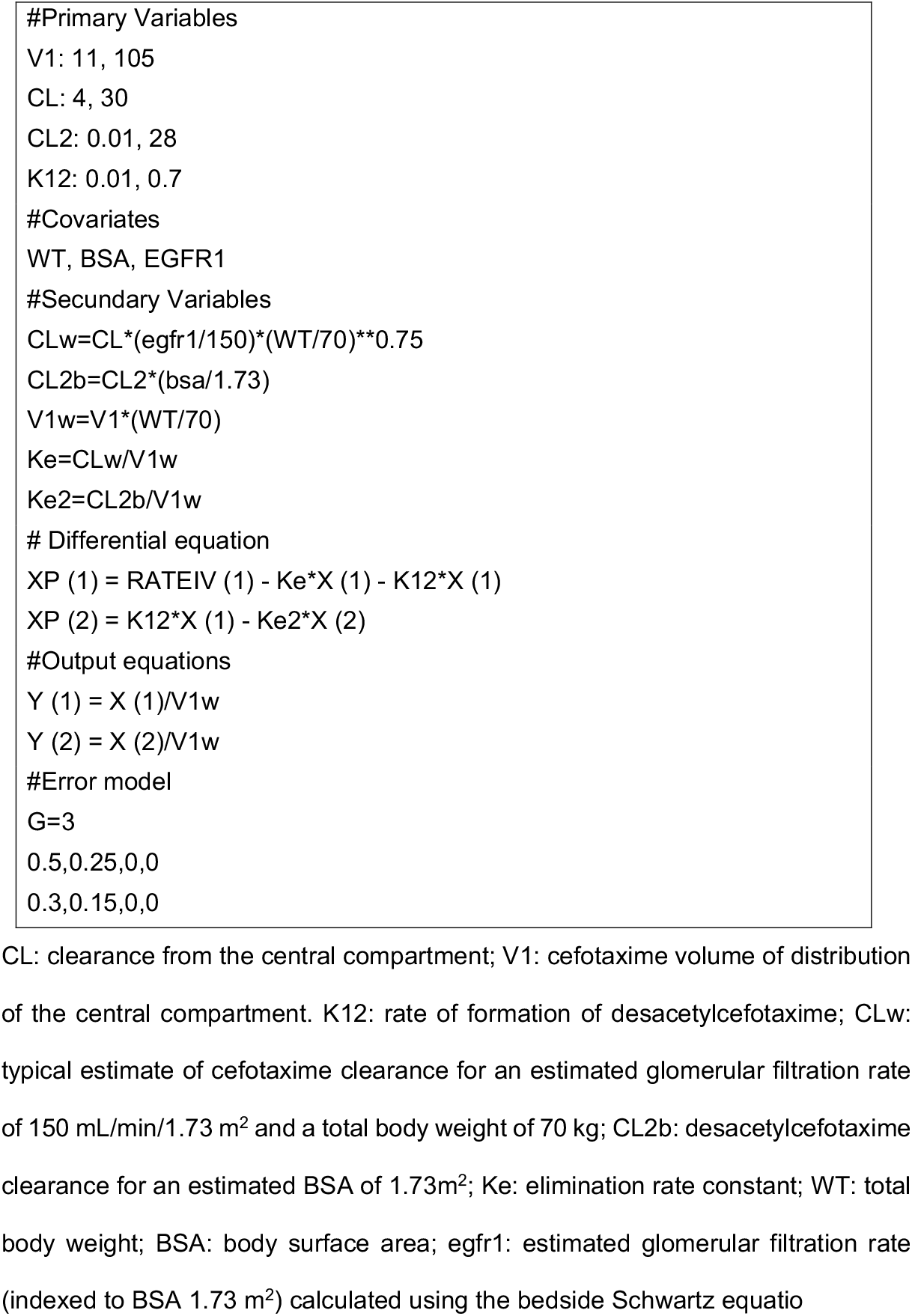
Model file used in Pmetrics for the final covariate model for cefotaxime and desacetylcefotaxime

**Figure S1:**
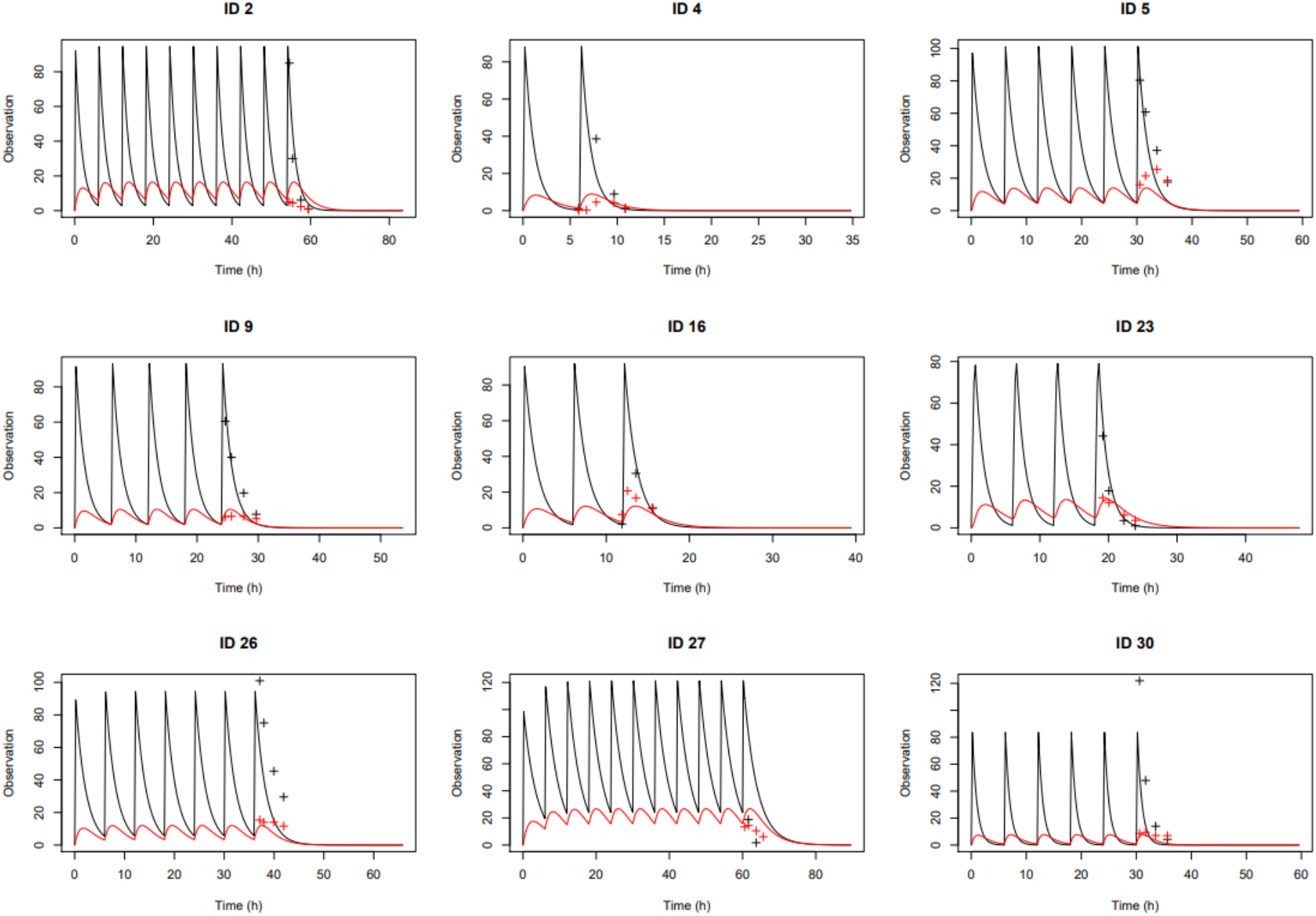
Individual plots for patients ID 2 to 30

**Figure S2:**
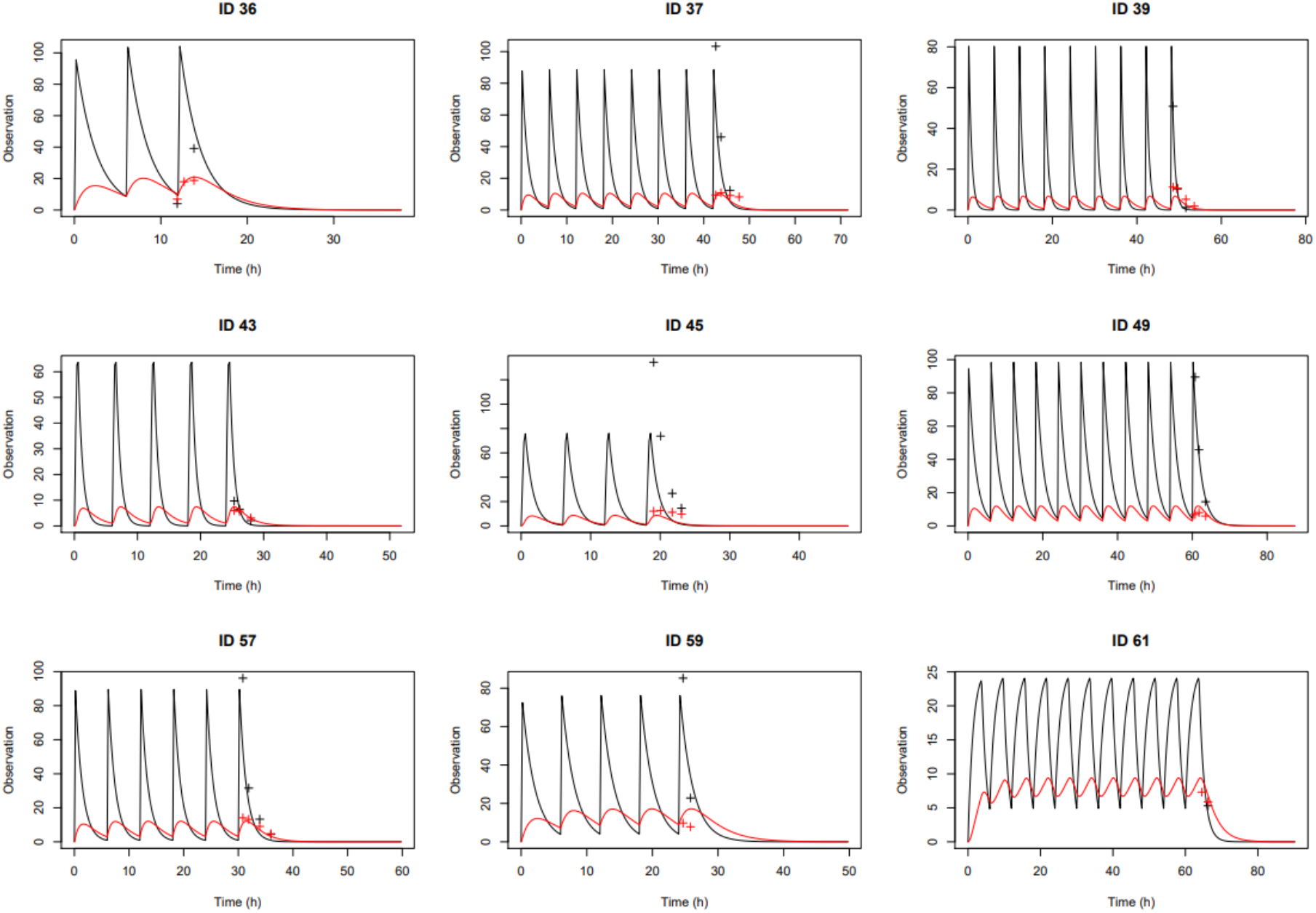
Individual plots for patients ID 36 to 61

**Figure S3:**
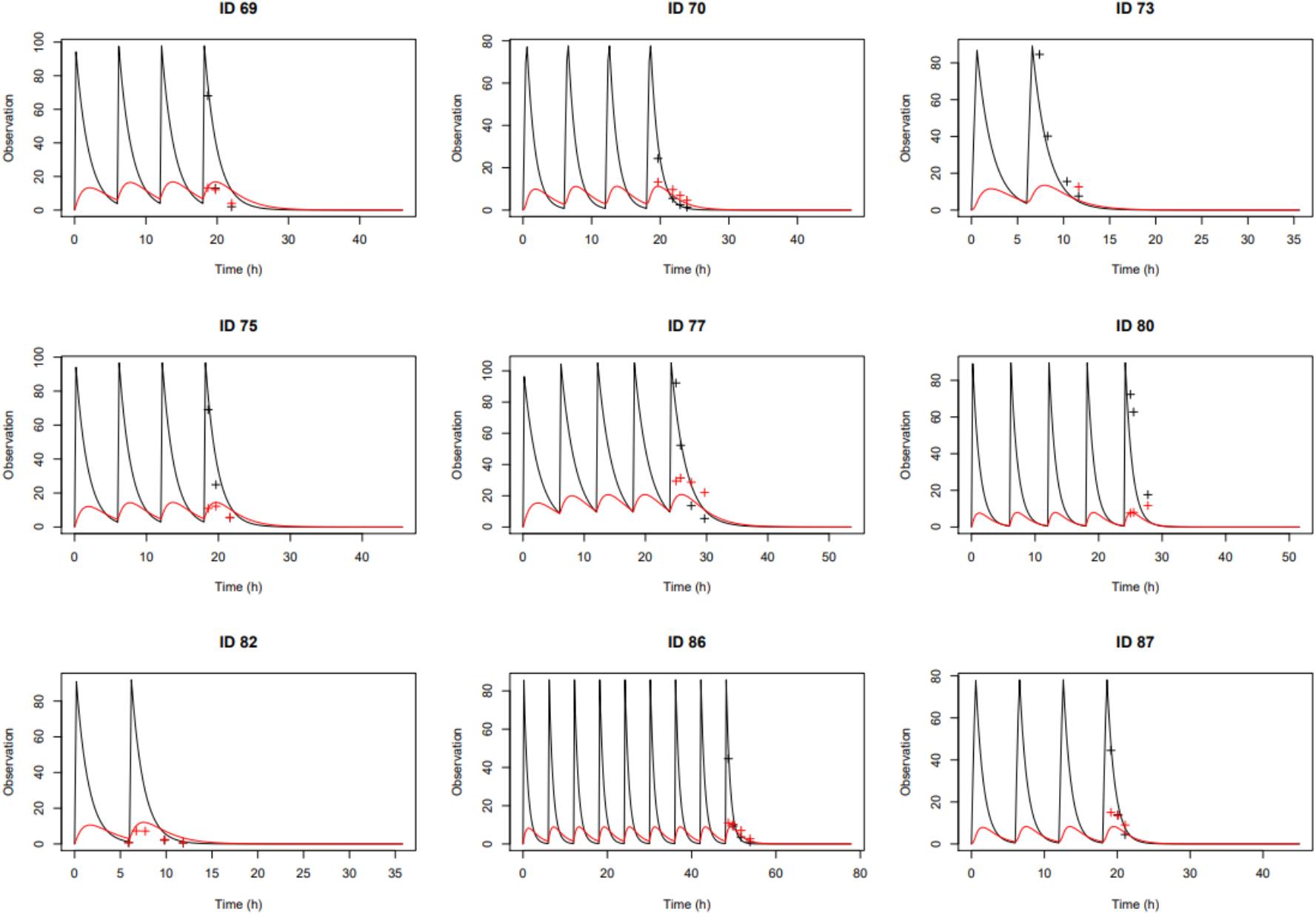
Individual plots for patients ID 69 to 87

**Figure S4:**
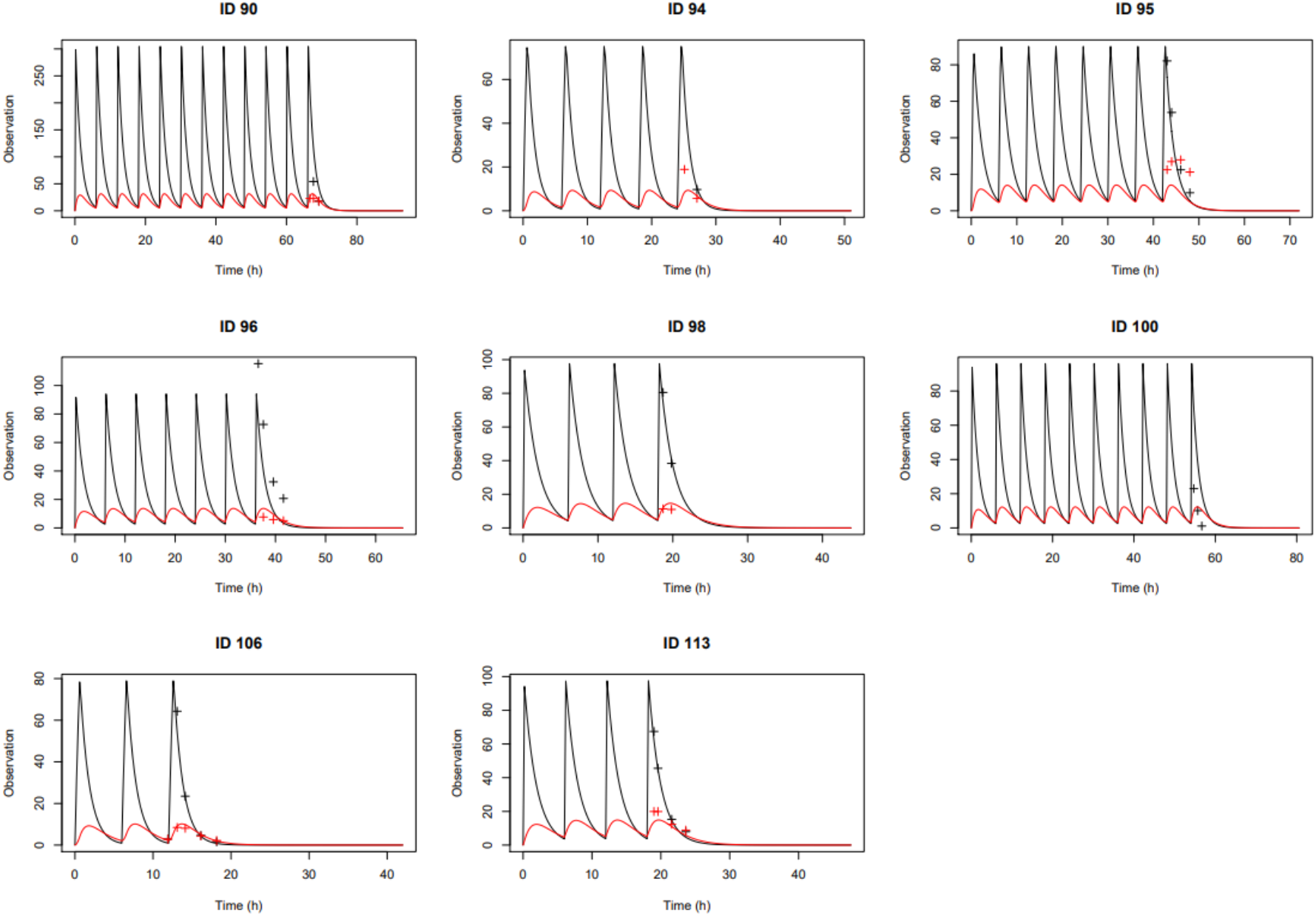
Individual plots for patients ID 90 to 113

## Notes

### Competing Interest Statement

The authors have declared no competing interest.

### Funding Statement

The study was funded by a grant from the Childrens Hospital Foundation, Queensland. Y Guerra Valero is a recipient of a Research Training Scholarship from The University of Queensland. SL Parker is a recipient of an Early Career Research Fellowship from the Australian National Health and Medical Research Council (APP1142757). JA Roberts is a recipient of an Australian National Health and Medical Research Council Fellowship (APP1048652).

### Author Declarations

Human Research and Ethics Committee of the Queensland Childrens Hospital gave ethical approval for this work

